# Face masks to control the source of respiratory infections: A systematic review of the scientific literature before and after COVID-19

**DOI:** 10.1101/2023.10.05.23296616

**Authors:** Elissa Brown, Alyson Haslam, Vinay Prasad

## Abstract

**Objective:** To examine the scientific literature on mask-use as source control to protect others from respiratory infections before and after the onset of the COVID-19 pandemic.

**Design:** Systematic review.

**Methods:** We examined primary research on mask usage as a means of source control to protect others by reducing the spread of respiratory diseases and contrasted the literature published before the onset of the COVID-19 pandemic with that published afterward. Articles were obtained through a search of PubMed and a review of article references. March 1, 2020 was selected as the cutoff date to distinguish between the pre-COVID-19 and post-COVID-19 periods.

**Results:** 195 articles met our inclusion criteria. The sample included 55 articles on source control published before the start of the COVID-19 pandemic, and 140 articles published after the pandemic began, representing a 154.5% increase. The percentage of randomized control trials (RCT) and cluster RCTs declined by 94.9% (p<0.001), representing only 1.4% of the post-pandemic literature. The percentage of studies conducted on human subjects declined by 48.8% (p<0.001), and the percentage of studies in healthcare facilities declined by 64.5% (p=0.019). One in 5 post-pandemic studies (21.4%) were conducted in “real world” settings; 1 in 10 post-pandemic studies (10.0%) were done with computer modeling. Study authors were significantly more supportive of masks as source control in the post-pandemic literature.

**Conclusions:** The quality of evidence in the published literature on masks as source control is lower after the start of the COVID-19 pandemic, with notable shifts in methodologies, research subjects, setting, and author tone.

## Introduction

Source control is a strategy for reducing respiratory disease transmission by blocking respiratory secretions that are produced through speaking, coughing, and sneezing. Simple hygiene measures like covering a cough or sneezing into a tissue are considered source control, as well as the use of face masks to prevent large respiratory droplets from traveling into the air and onto other people. Recommending masks as a means of source control to slow the spread of SARS-CoV-2 infection became a topic of urgent importance during the COVID-19 pandemic, due to the possibility of asymptomatic and pre-symptomatic transmission.^1^ While early studies raised concern that a significant portion of individuals with COVID-19 lacked symptoms but could still transmit the virus to others, later studies showed that asymptomatic individuals who tested positive for SARS-CoV-2 were far less likely to be infectious than initially believed.^2, 3^ Official masking guidance and policies were enacted in many contexts beginning in March 2020, based on the premise that a large portion of healthy people may unknowingly be infected and could transmit the virus to others. Thirty-nine states, Puerto Rico and the District of Columbia issued orders requiring people to wear masks outside the home in 2020.^4^ Partisan debate over mask mandates became a central feature of the pandemic in the United States. Uncertainty around mask efficacy was downplayed by public health experts, attempts at open discussion and scientific debate were undermined, and dissenting viewpoints were flagged as misinformation.^5^ Ultimately the United States became an international outlier with the CDC recommending masks for anyone over two-years of age, and some states requiring school children to mask well into 2022.^6^

The CDC maintains guidelines for universal masking and emphasizes the role of source control for respiratory disease transmission, stating that “masks contain droplets and particles you breathe, cough, or sneeze out so you do not spread them to others”.^7^ In the community setting, the CDC continues to recommends anyone ages two-years or older should wear masks in indoor public spaces when the COVID-19 hospital admission level is high.^8^ In healthcare settings, they recommend masks as source control for ten days after a known COVID-19 exposure, and recommend universal masking when a facility is experiencing a SARS-CoV-2 or other outbreak of respiratory infection.^9^ In the Fall of 2023, universal mask mandates were reinstated in a number of institutions in the U.S. including several colleges and universities, along with large hospital systems in California and New York.^10^ The ongoing implementation of mask mandates continues to prompt debate as to whether these policies are supported by evidence.^6, 11, 12, 13^

Many observational and ecologic studies on mask use and mask mandates have been published since the COVID-19 pandemic began. While the body of published research on masks has grown, it is unknown to what degree these changes are the result of new literature on masks used as source control. It is with this background that we conducted a systematic review of the scientific literature pertaining to mask-use as source control for respiratory infections, and compared findings from before and after the onset of the COVID-19 pandemic.

## Methods

We reviewed the scientific literature on masks used as source control and compared the findings from before and after the onset of the COVID-19 pandemic to observe whether any significant changes occurred. For the purposes of this analysis, we designate literature published on or before February 29, 2020 as occurring before the pandemic and anything published on or after March 1, 2020 as post-pandemic literature.

### Article Selection

A PubMed search was conducted on February 2, 2023 with the search phrase “masks AND ‘source control’ AND infection”. Source control is defined as evidence that mask wearing provides a benefit to third parties in and around the masking person. No study designs were excluded. Included articles were original research articles, referred to the intervention as a “source control”, or considered masks for source control as part of universal masking protocol. Articles published before the search date were selected for analysis based on relevance to masking as source control; for example, simulations where masks were placed on “source” mannequins, intervention trials where index (“source”) patients were randomized to mask around healthy family members, and pre-post studies measuring transmission after universal masking policies were enacted, with the public health rationale for these mandates largely attributed to masks preventing spread of COVID-19 from asymptomatic sources. Similarly, mask intervention studies where all participants (“source” and “receiver”) were instructed to wear masks, and ecological/observational studies on the impact of mask mandates in various settings (e.g. hospital, dormitory, household) were deemed relevant. Studies were excluded if index (“source”) cases were not masked, such as sick children who were not instructed to mask while caregivers or healthcare workers wore masks for their own protection. Similarly, articles were excluded if masks were considered only as personal protective equipment (PPE) with no source control component, as well as articles with endpoints specific to masking as personal protection only, such as secondary transmission rates among masked hospital workers. Articles pertaining to source control in the operating room or during procedures to prevent post-surgical wound infection were excluded. Moreover, we excluded articles that were not in English, and articles examining mask use in situations unrelated to respiratory infection, such as chemical pollutant exposure or airplane filtration.

Finally, we reviewed the references of studies that met our inclusion criteria to identify any additional relevant studies for inclusion, due to the fact that mask research in the post-COVID time frame has focused heavily on universal masking to prevent the spread of SARS-CoV-2 with source control as one component. The term “source control” is infrequently used when referring to COVID-19 universal masking protocol in this body of literature. For example, many of the articles the CDC references in support of universal masking to control the source of SARS-CoV-2 infection do not consistently utilize the term “source control”.^14^ By including these additional articles we feel we have most comprehensively captured the available literature on source control despite variations in terminology.

### Data Abstraction

For each article we abstracted the following data: title, author(s), journal, date of publication, study design, geographic location of primary investigator, research setting (including healthcare, community or laboratory), research population (human subjects vs. mannequins), mask type(s), and results/findings.

### Statistical Analysis

The data were divided into two groups based on publication date: 1) Studies published prior to March 1, 2020, which represents the pre-pandemic time period, and 2) Studies published from March 1, 2020 to February 6, 2023 (date of search), which represents the post-pandemic time period. Because we used publicly available data, which did not include personally identifiable information, an institutional review board’s approval was not required.

For each time period we determined the total number of articles relevant to source control. We then categorized the articles by study design, including: RCT/cluster RCT, pre/post-intervention (with no separate control group), observational studies (cohort, cross-sectional, case-control, ecological), laboratory experiment (human participants, mannequin or other simulation equipment), computer modeling, reviews, and commentary. We calculated the proportion of articles for each study type as a percent of the total for each time period and then calculated relative changes in the proportion of each type of study.

We categorized articles by the research setting, including: healthcare setting, community setting, laboratory setting and no setting (e.g. review articles, commentary, computer modeling studies). Articles were also categorized by study population, including: human subject, animal subject, mannequins or other simulation equipment, and no subject (e.g. review articles, commentary, computer modeling studies). We then calculated the proportion of articles conducted in each setting in both time periods and for each study population in each time period, and determined whether any of the changes in proportion were significant. Finally, the full text articles (n=179) were reviewed for author tone by two raters, with each article receiving a rating of supportive of masks, unsupportive of masks, or equivocal. The proportion of articles with each tone were calculated in both time periods and compared to determine whether any of the changes in proportion were significant.

We accepted a p-value of 0.05 to determine if the differences in proportions before and after the pandemic were statistically significant (see Table 1). We used R statistical software (version 4.2.1) [9] to conduct data analysis. For tone, we validated interrater agreement by calculating agreement proportions and kappa statistics using R statistical software (version 4.2.1) [9] with package ‘irr’.

**Table 1:** Comparison of source control literature published before and after the start of the COVID-19 pandemic by study type, study setting, study population and tone.

|  | Before Pandemic N (%) | After Pandemic N (%) | Relative Percent Change | p-value |
| --- | --- | --- | --- | --- |
| ***Type of Study (n=195)*** |  |  |  |  |
| Lab Experiment | 17 (30.9) | 39 (27.9) | -9.7% | 0.80 |
| RCT/Cluster RCT | 15 (27.3) | 2 (1.4) | -94.9% | **< 0.001** |
| Review | 13 (23.6) | 47 (33.6) | 42.4% | 0.24 |
| Pre/Post Intervention | 6 (10.9) | 11 (7.9) | -27.5% | 0.69 |
| Observational | 2 (3.6) | 17 (12.1) | 236.10% | 0.13 |
| Computer Modeling | 1 (1.8) | 14 (10.0) | 454.6% | 0.10 |
| Commentary | 1 (1.8) | 10 (7.1) | 294.4% | 0.27 |
| ***Study Setting (n=195)*** |  |  |  |  |
| Laboratory | 17 (30.9) | 39 (27.9) | -9.7% | 0.80 |
| Healthcare | 11 (20.0) | 10 (7.1) | -64.5% | **0.019** |
| Community | 12 (21.8) | 20 (14.3) | -34.4% | 0.29 |
| No setting | 15 (27.3) | 71 (50.7) | 85.7% | **0.005** |
| ***Study Population (n=195)\**** |  |  |  |  |
| Human | 30 (54.5) | 39 (27.9) | -48.8% | **< 0.001** |
| Mannequins | 8 (14.5) | 31 (22.1) | 52.4% | 0.32 |
| Animal | 0 (0.0) | 1 (0.7) | -- | 1.0 |
| None | 17 (30.9) | 71 (50.7) | 64.1% | **0.019** |
| ***Author Tone (n=179)\*\**** |  |  |  |  |
| Supportive | 28 (60.9) | 114 (85.7) | 40.7% | **< 0.001** |
| Unsupportive | 11 (23.9) | 8 (6.0) | -74.9% | **0.002** |
| Equivocal | 7 (15.2) | 11 (8.3) | -45.4% | 0.29 |

\*Cumulative proportion post-COVID exceeds 100%, as two studies incorporated both human and mannequin subjects; mannequin category includes all simulation equipment.
\*\*Assessed for all articles with full text access.

## Results

A total of 389 articles were identified in the literature search. An initial title screen resulted in the exclusion of 105 for non-English language and for non-relevance; 107 additional articles were excluded for relevance after screening abstracts (e.g. non-respiratory infection, chemical pollutant, airplane filtration). The remaining 177 articles were read in full and 92 were determined to fit our inclusion criteria. A thorough review of references yielded 103 additional articles for inclusion, which were consistent with the concept of source control but did not appear in the original search primarily due to their focus on universal masking. The final analytic sample includes 195 scientific articles published on masks as source control (Supplement).

Using our search strategy, we identified 55 scientific articles published on source control prior to the start of the COVID-19 pandemic, and 140 articles published after the pandemic began, representing a 154.5% increase in articles published on source control (Figure 1). Randomized control trials (RCT) and cluster RCTs comprised 27.3% of the pre-pandemic total (15/55) and 1.4% of the post-pandemic total (2/140), representing a -94.8% relative change in RCTs/cluster RCTs (p<0.001). Relative changes in proportion were observed for all other study types, including lab experiments (-9.7%), reviews (42.4%), pre/post interventions (-27.5%), observational studies (236.1%), computer modeling studies (454.6%) and commentary (294.4%); however, these findings were not significant at p=0.05. (Table 1).

**Figure 1:**
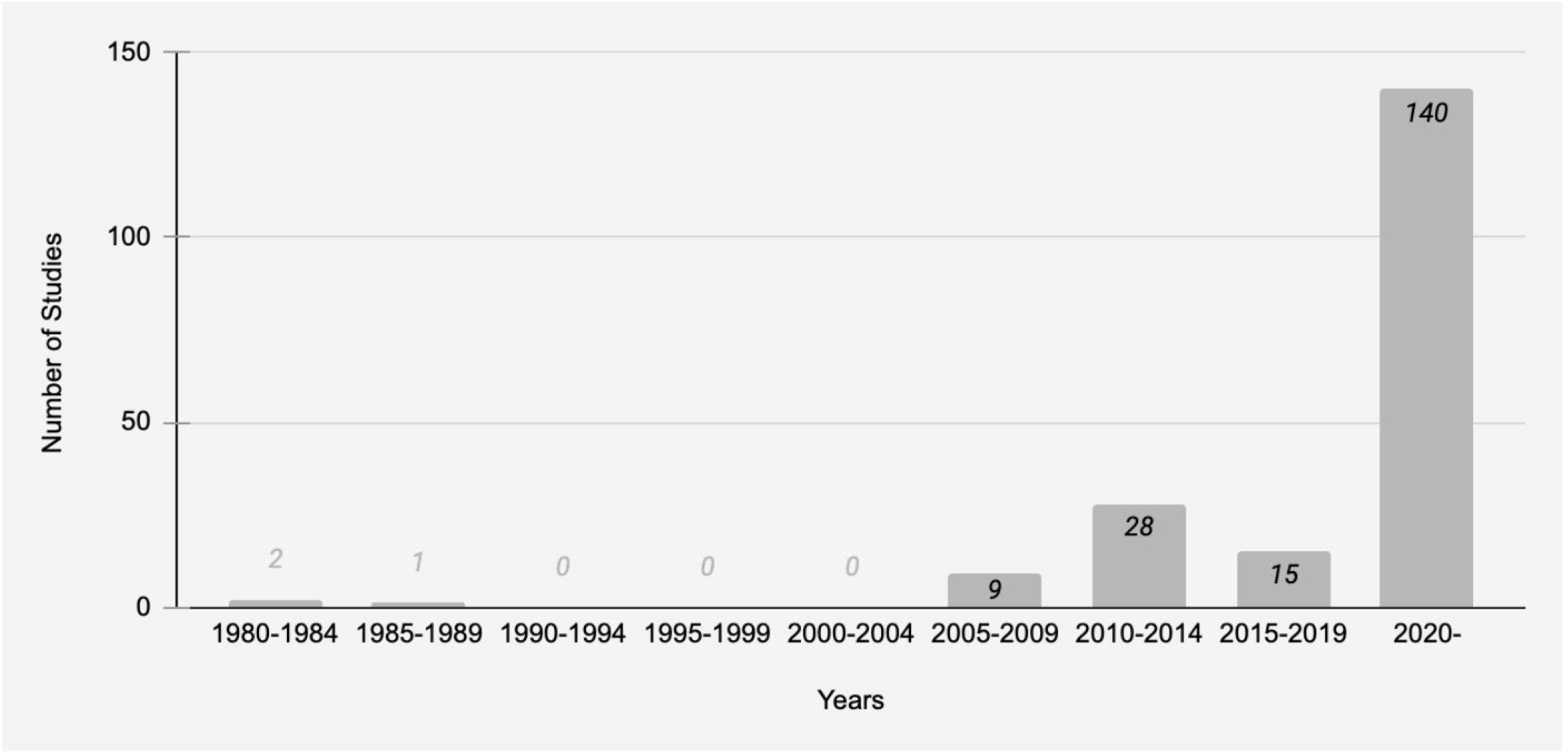
Scientific studies published on face masks as source control to protect others.

Laboratory experiments were the most prevalent study type published on source control before the pandemic, comprising 30.9% of the published literature (17/55); reviews were the most prevalent study type in the post-pandemic time period, comprising 33.6% percent of the post-pandemic total (47/140). RCTs were the second most prevalent study type pre-pandemic (15/55) representing 27.3% of the total, and the least prominent study type in the post-pandemic period at 1.4% (2/140). Study types ranked from most to least prevalent prior to the COVID-19 pandemic are: 1. Lab Experiments (n=17), 2. RCTs/Cluster RCTs (n=15), 3. Reviews (n=13), 4. Pre/post-Intervention (n=6), 5. Observational study (n=2), 6. Computer modeling (n=1), and 7. Commentary (n=1). The study types ranked from most to least prevalent in the time period after the pandemic began are: 1. Reviews (n=47), 2. Lab Experiments (n=39), 3. Observational (n=17), 4. Computer modeling (n=14), 5. Pre/post-Intervention (n=11), 6. Commentary (n=10), 7. RCT/cluster RCT (n=2). (Figure 2).

**Figure 2:**
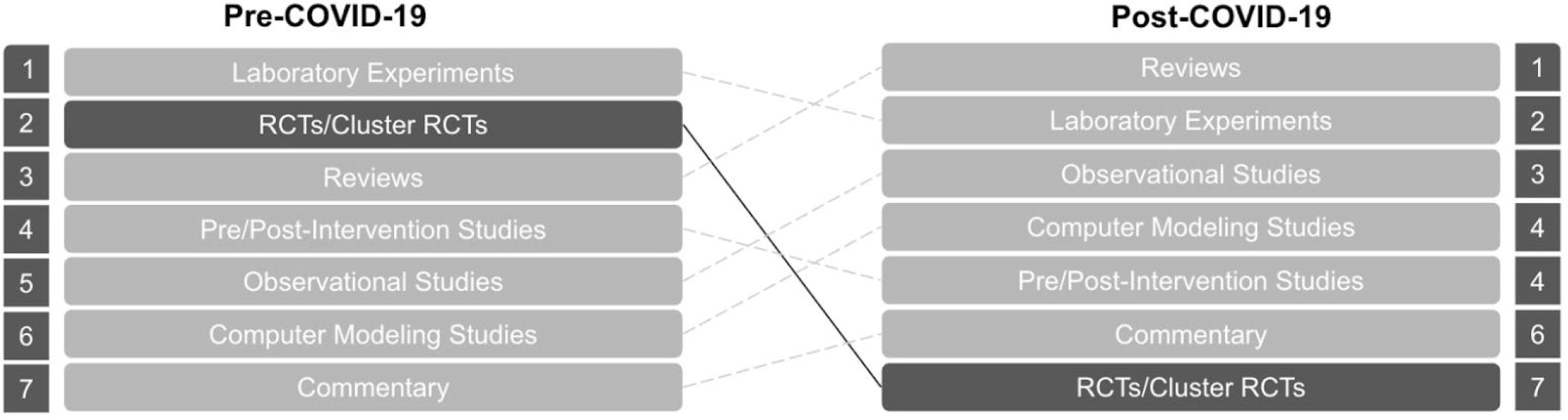
Study types ranked by percentage of total published literature on face masks as source control before and after the start of the COVID-19 pandemic

Studies conducted in healthcare settings make up 20.0% of the total (11/55) prior to the pandemic, and represent 7.1% of the total after the pandemic began (10/140), a relative change of -64.5% (p=0.019). The percentage of studies without a setting (e.g. computer modeling, reviews) increased by 85.7%, from 27.3% before the pandemic (15/55) to 50.7% after the pandemic began (72/140) (p=0.005). The percentage of studies conducted in community settings and laboratory settings declined by 34.4% and 9.7%, respectively, but these changes were not statistically significant at the p=0.05 level. (Table 1, Figure 3).

**Figure 3:**
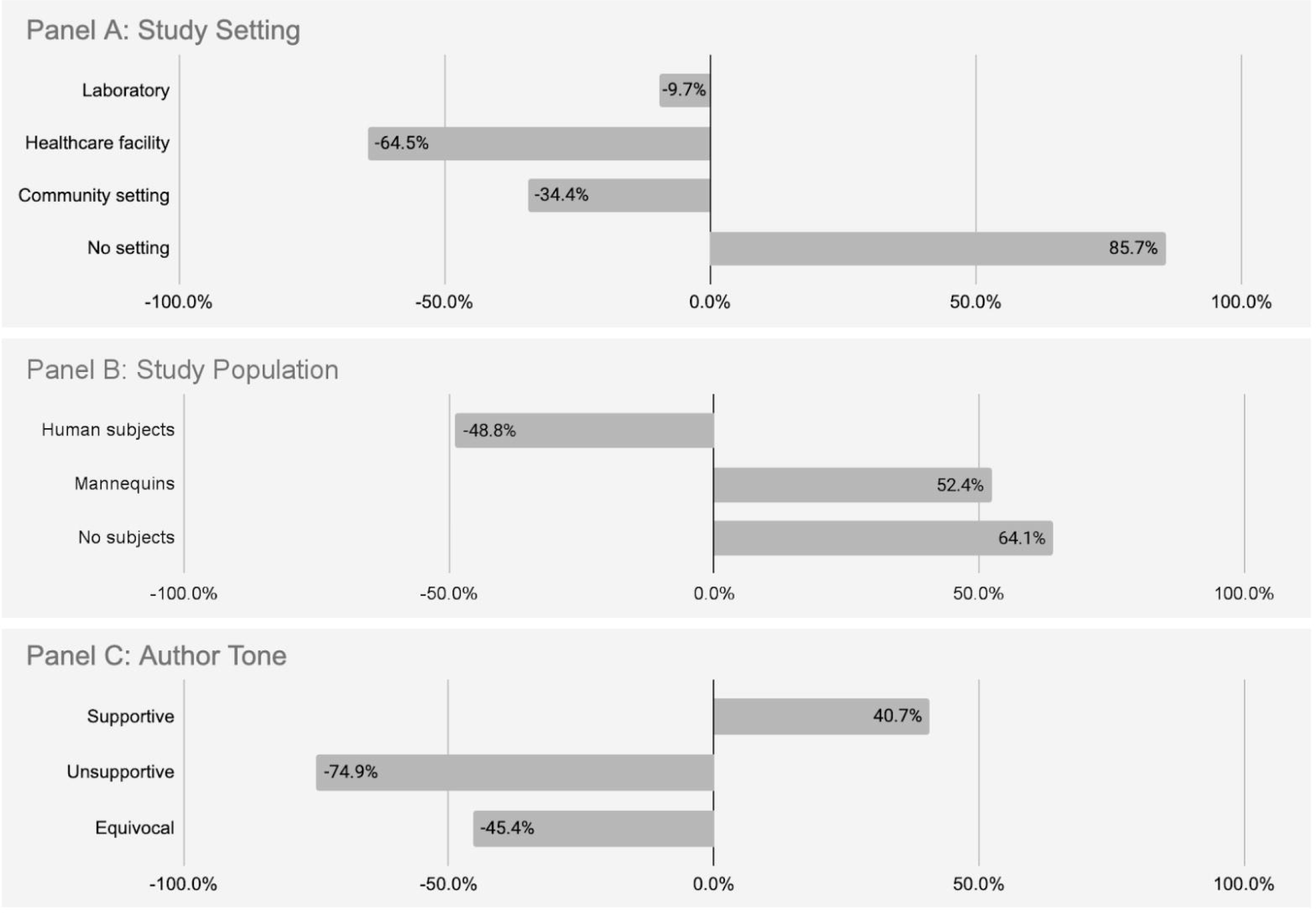
Changes in study setting (Panel A), study population (Panel B), and author tone (Panel C) for face masks as source control published before and after the start of the COVID-19 pandemic

More than half (54.5%) of studies published before the pandemic involved human subjects (30/55) and 27.9% of post-pandemic studies involved human subjects (39/140), representing a -48.8% relative change in the proportion of studies conducted using human subjects (p<0.001). Studies with no subjects (e.g. computer modeling studies, review articles, commentary) increased by 64.1%, from 30.9% of pre-pandemic studies (17/55) to 50.7% of post-pandemic studies (71/140) (p=0.019). The proportion of studies involving mannequins and other simulation equipment increased by 52.4%, but this change was not significant at the p=0.05 level. (Table 1, Figure 3).

The percentage of articles that were supportive toward masks as source control increased by 40.7% (p<0.001) from the pre- to post-pandemic study period, from 60.9% (28/46) to 85.7% (114/133) of all published studies. The percentage of articles that were unsupportive of masks as source control declined by 74.9% (p=0.002), from 23.9% (11/36) pre-pandemic to just 6.0% (8/133) of articles in the post pandemic study period. (Table 4). Inter-rater agreement was assessed using Cohen’s Kappa statistic, and measured to be 0.935 (p<0.001), indicating strong agreement. (Table 1, Figure 3).

## Discussion

The proliferation of studies on masks as source control since March 2020 parallels the dramatic increase in the utilization of this particular public health measure due to pandemic response efforts like universal mask mandates, which were largely based on source control to prevent asymptomatic spread. Our research indicates that the quality of evidence in the published literature on masks as source control declined over the course of the COVID-19 pandemic, with notable shifts in methodologies, research subjects, setting and author tone. Prior to the COVID-19 pandemic, randomized controlled trials (RCTs) and cluster RCTs were the second most prevalent type of study on masks as source control, yet comprise only 1.4% of the total published literature on source control in the post-pandemic study period (n=2).

While this shift reflects the practical challenges associated with conducting RCTs during a pandemic, a gold-standard of evidence must be upheld when public health policies and mandates continue long after a pandemic state of emergency has ended. In this case, both RCTs conducted in the post-COVID timeframe were large-scale cluster trials of universal masking protocol with source control as one component, and neither study found a significant impact of masks on viral spread. The first published by Alfelali et al in 2020 allocated patients at three consecutive Hajj pilgrimages to “face mask” (universal masking protocol) or “no face mask” (control) groups, and was unable to provide conclusive evidence on facemask efficacy against viral respiratory infections.^14^ A second cluster RCT was published by Abaluck et al in 2022, which reported decreases in both symptomatic seroprevalence and reported COVID-19-like symptoms, but the effect size was small and there was a lack of statistical significance.^15^ A subsequent reanalysis by Chikina et al notes the presence of unblinding, ascertainment bias, and bias-susceptible endpoints, which warrant caution and complicate drawing any causal link between masks and the small observed decrease in population-rate of symptomatic seropositivity; while the mask intervention was highly effective at modifying behaviors (distancing, mask-wearing, symptom reporting), the data on mask wearing was consistent with having modest or no direct effect on COVID-related outcomes.^16^

Our research also found that the majority of studies on source control published post-pandemic (n=101, 72.1%) were conducted with no human subjects, and just 1 in 5 were conducted in “real world” settings such as in healthcare facilities or in the community. A scant 1.8% of published studies on source control before the pandemic relied on computer modeling, while 1 out of every 10 studies published on source control post-pandemic utilized computer modeling. Again, this shift reflects a number of practical and ethical challenges to conducting research during a pandemic, but the lack of studies conducted on humans in real world settings should be considered when examining the overall body of evidence and setting long term public health policy.

Finally, the studies published during the post-pandemic study period present as significantly more favorable toward masking as source control than prior to the pandemic, with only 6% of published literature in the post-pandemic study period representing an unsupportive tone compared to 86% supportive toward masks. This shift in tone may be a reflection of pandemic-related selection bias from journals and may not be representative of all studies that were submitted for publication. Indeed, in a previous examination of the evidence for facemask efficacy published in the MMWR, which represents a fraction of the published mask efficacy studies, there were no studies published on the topic prior to the COVID-19 pandemic, and most of the published MMWR studies used causal language about the effectiveness of masks despite only a small percentage actually testing the masks.^17^

### Limitations

Our study has several limitations. We did not include non-English language publications or studies in this research. We attempted to compile an exhaustive list of published studies on masking as source control, but there may be other articles in the body of literature as no single convention exists for referring to this subject. Related is that the term “source control” is not used consistently from study to study, but for the purposes of this analysis, we applied a uniform a priori definition in determining included and excluded articles.

## Conclusions

Maintaining research quality and rigor is essential to informing effective public health measures during a pandemic and beyond. Limitations in the quality of new evidence on masks as source control during and after the COVID-19 pandemic may be attributed to the urgency of the situation, yet should be weighed accordingly when using this body of literature to inform ongoing public health policy.

The trend toward relying on non-randomized, non-real-world, and even non-human studies in the post-COVID-19 literature is a concern. Randomized studies, especially when there is equipoise on a topic, are essential in determining the efficacy of an intervention.^11^ Whether masks can be used as source control has been debated at length, and the inconsistent recommendations by different government health agencies underscores the need for high-quality data on this topic.

Finally, recent findings challenge the very premise of using masks to control the source of asymptomatic spread, which is the justification for universal masking and mask mandates. Asymptomatic individuals who tested positive for SARS-CoV-2 upon hospital admission screening were far less likely to be infectious than previously believed. The vast majority of patients (96%) found to be SARS-CoV-2 positive by PCRs on hospital admission were non-infectious based on strand-specific testing.^3^ This finding has profound implications for public health policies, and should be given significant weight when assessing the role of universal masking and mask mandates as source control for mitigating the spread of COVID-19 and other respiratory illnesses.

## Supporting information

Supplement: Article selection diagram

## Data Availability

All data produced are available online at Pubmed.

## References

(1) Bai Y, Yao L, Wei T, et al. Presumed Asymptomatic Carrier Transmission of COVID-19. JAMA. 2020;323(14):1406–1407. doi:10.1001/jama.2020.2565

(2) Methi, F., Madslien, E.H. Lower transmissibility of SARS-CoV-2 among asymptomatic cases: evidence from contact tracing data in Oslo, Norway. BMC Med 20, 427 (2022). 10.1186/s12916-022-02642-4

(3) Tayyar, R., Kiener, M., Liang, J., Contreras, G., Rodriguez-Nava, G., Zimmet, A., … Salinas, J. (2023). Low infectivity among asymptomatic patients with a positive severe acute respiratory coronavirus virus 2 (SARS-CoV-2) admission test at a tertiary care center, 2020–2022. Infection Control & Hospital Epidemiology, 1–3. doi:10.1017/ice.2023.210

(4) Markowitz, Andy. State-by-State Guide to Face Mask Requirements. AARP. Updated September 28, 2023. Accessed September 28, 2023 from: https://www.aarp.org/health/healthy-living/info-2020/states-mask-mandates-coronavirus.html.

(5) Russell, J.H and Patterson, D. The Mask Debacle: How partisan warfare over mandates became a central feature of the pandemic. Tablet Magazine. Published February 16, 2022. Accessed October 1, 2023. https://www.tabletmag.com/sections/science/articles/the-mask-debacle

(6) Høeg TB, González-Dambrauskas S, Prasad V. The United States’ decision to mask children as young as two for COVID-19 has been extended into 2023 and beyond: The implications of this policy. Paediatr Respir Rev. 2023;47:30–32. doi:10.1016/j.prrv.2023.04.004

(7) Centers for Disease Control and Prevention. How to Protect Yourself and Others. Updated July 6, 2023. Accessed on October 3, 2023 from: https://www.cdc.gov/coronavirus/2019-ncov/prevent-getting-sick/prevention.html

(8) Centers for Disease Control and Prevention. COVID-19 by County. Updated May 11, 2023. Accessed October 3, 2023 from: https://www.cdc.gov/coronavirus/2019-ncov/your-health/covid-by-county.html

(9) Centers for Disease Control and Prevention. Interim Infection Prevention and Control Recommendations for Healthcare Personnel During the Coronavirus Disease 2019 (COVID-19) Pandemic. Updated May 3, 2023. Access on October 3, 2023 from: https://www.cdc.gov/coronavirus/2019-ncov/hcp/infection-control-recommendations.html

(10) Rahman, K. Mask Mandates Return: Full List of Places With Restrictions in Place. Newsweek. Published on August 29, 2023. Accessed on October 3, 2023 from: https://www.newsweek.com/mask-mandates-return-list-places-restrictions-1823069

(11) Høeg TB, González-Dambrauskas S, Prasad V. Does equipoise exist for masking children for COVID-19?Public Health Pract (Oxf). 2023 Sep 9;6:100428. doi: 10.1016/j.puhip.2023.100428. PMID: 37744300; PMCID: PMC10511791.

(12) Brown E, Haslam A, Prasad V. Flu advice in the U.S. news media changed during the COVID-19 pandemic but not the evidence. European Journal of Epidemiology. ublished online 2023:1–3. doi:10.1007/s10654-023-01037-w

(13) Miller, S. and Prasad, V. Changes in Masking Policies in US Healthcare Facilities in the First Quarter of 2023: Do COVID-19 Cases, Hospitalizations, or Local Political Preferences Predict Loosening Restrictions. Posted July 12, 2023. Access on October 3, 2023 from: https://www.medrxiv.org/content/10.1101/2023.07.11.23292518v1

(14) Centers for Disease Control and Prevention. The Science of Masking to Control COVID-19. Published November 16, 2020. Accessed on October 3, 2023 from: https://www.cdc.gov/coronavirus/2019-ncov/downloads/science-of-masking-abbreviated.pdf

(15) Alfelali M, Haworth EA, Barasheed O, Badahdah AM, Bokhary H, Tashani M, Azeem MI, Kok J, Taylor J, Barnes EH, El Bashir H, Khandaker G, Holmes EC, Dwyer DE, Heron LG, Wilson GJ, Booy R, Rashid H; Hajj Research Team. Facemask against viral respiratory infections among Hajj pilgrims: A challenging cluster-randomized trial. PLoS One. 2020 Oct 13;15(10):e0240287. doi: 10.1371/journal.pone.0240287. PMID: 33048964; PMCID: PMC7553311.

(16) Abaluck J, Kwong LH, Styczynski A, et al. Impact of community masking on COVID-19: A cluster-randomized trial in Bangladesh. Science. 2022;375(6577):eabi9069. doi:10.1126/science.abi9069

(17) Chikina M, Pegden W, Recht B. Re-analysis on the statistical sampling biases of a mask promotion trial in Bangladesh: a statistical replication. Trials. 2022 Sep 15;23(1):786. doi: 10.1186/s13063-022-06704-z. PMID: 36109816; PMCID: PMC9479361.

(18) Høeg TB, Haslam A, Prasad V. An analysis of studies pertaining to masks in Morbidity and Mortality Weekly Report: Characteristics and quality of studies from 1978 to 2023. Am J Med. 2023 Sep 28:S0002–9343(23)00580-6. doi: 10.1016/j.amjmed.2023.08.026. Epub ahead of print. PMID: 37777144.

