## Supplementary figures and images for "Face masks to control the source of respiratory infections: A systematic review of the scientific literature before and after COVID-19"

### Supplement: Article selection diagram

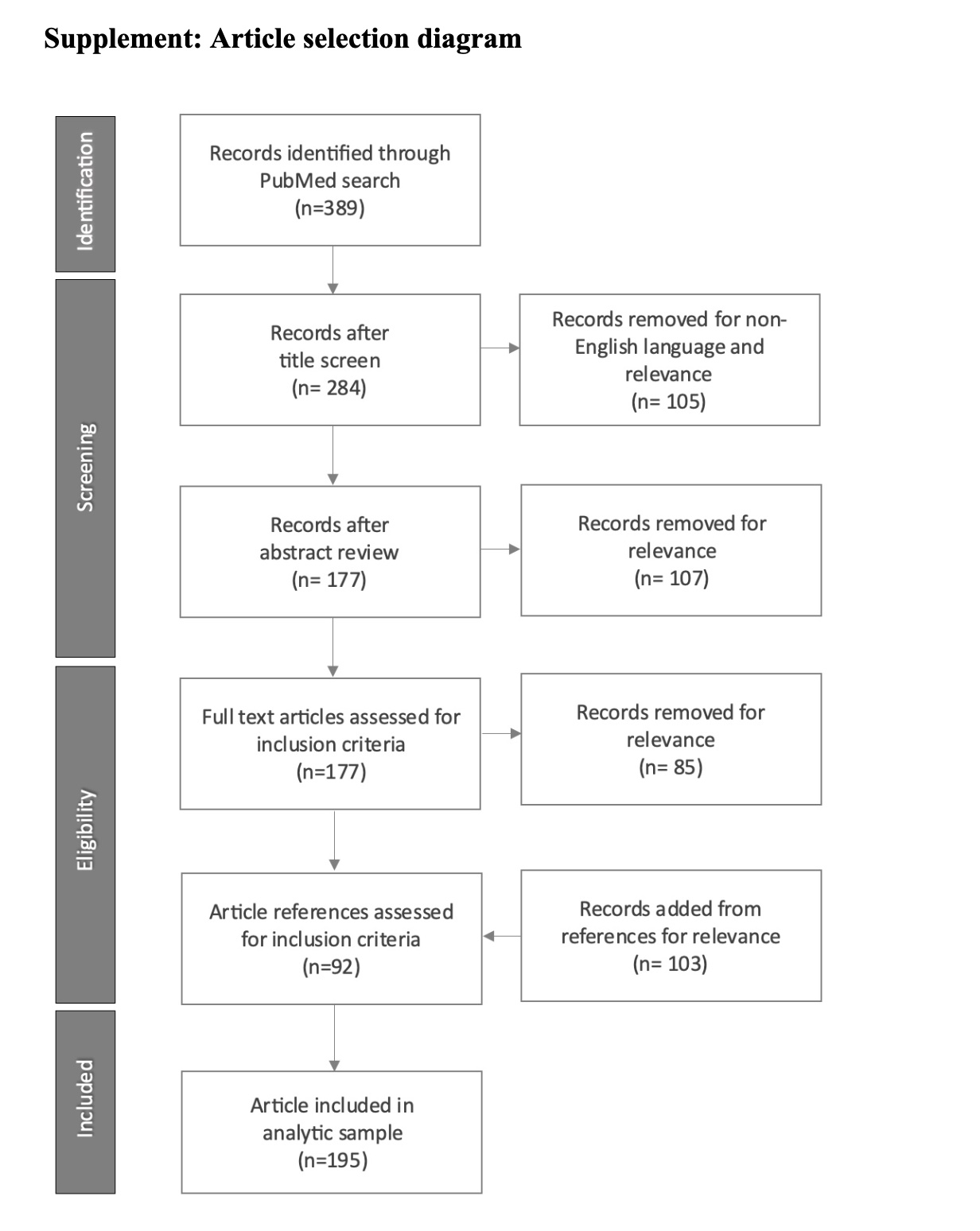
